# A deep learning model for clinical outcome prediction using longitudinal inpatient electronic health records

**DOI:** 10.1101/2025.01.21.25320916

**Authors:** Ruichen Rong, Zifan Gu, Hongyin Lai, Tanna L. Nelson, Tony Keller, Clark Walker, Kevin W. Jin, Catherine Chen, Ann Marie Navar, Ferdinand Velasco, Eric D. Peterson, Guanghua Xiao, Donghan M. Yang, Yang Xie

**Author notes:** Equal contribution as first authors (Ruichen Rong, Zifan Gu). Co-corresponding authors: Donghan M. Yang, PhD, The University of Texas Southwestern Medical Center Danciger Research Building, 5323 Harry Hines Blvd. Ste H9.124 Dallas, TX 75390-8821, Yang Xie, PhD, The University of Texas Southwestern Medical Center Danciger Research Building, 5323 Harry Hines Blvd. Ste H9.124 Dallas, TX 75390-8821.

## Abstract

**Objective:** Recent advances in deep learning show significant potential in analyzing continuous monitoring electronic health records (EHR) data for clinical outcome prediction. We aim to develop a Transformer-based, Encounter-level Clinical Outcome (TECO) model to predict mortality in the intensive care unit (ICU) using inpatient EHR data.

**Materials and Methods:** TECO was developed using multiple baseline and time-dependent clinical variables from 2579 hospitalized COVID-19 patients to predict ICU mortality, and was validated externally in an ARDS cohort (n=2799) and a sepsis cohort (n=6622) from the Medical Information Mart for Intensive Care (MIMIC)-IV. Model performance was evaluated based on area under the receiver operating characteristic (AUC) and compared with Epic Deterioration Index (EDI), random forest (RF), and extreme gradient boosting (XGBoost).

**Results:** In the COVID-19 development dataset, TECO achieved higher AUC (0.89–0.97) across various time intervals compared to EDI (0.86–0.95), RF (0.87–0.96), and XGBoost (0.88–0.96). In the two MIMIC testing datasets (EDI not available), TECO yielded higher AUC (0.65–0.76) than RF (0.57–0.73) and XGBoost (0.57–0.73). In addition, TECO was able to identify clinically interpretable features that were correlated with the outcome.

**Discussion:** TECO outperformed proprietary metrics and conventional machine learning models in predicting ICU mortality among COVID-19 and non-COVID-19 patients.

**Conclusions:** TECO demonstrates a strong capability for predicting ICU mortality using continuous monitoring data. While further validation is needed, TECO has the potential to serve as a powerful early warning tool across various diseases in inpatient settings.

**LAY SUMMARY:** In intensive care units (ICUs), accurately estimating the risk of death is crucial for timely and effective medical intervention. This study developed a new AI algorithm, TECO (Transformer-based, Encounter-level Clinical Outcome model), which uses electronic health records to continuously predict ICU mortality after admission, with the capability to update predictions on an hourly basis. TECO was trained on data from over 2,500 COVID-19 patients and was designed to analyze multiple types of continuous monitoring data collected during a patient’s ICU stay. We tested TECO’s performance against a widely used proprietary tool, the Epic Deterioration Index (EDI), and other machine learning methods, such as random forest and XGBoost, across three patient groups: COVID-19, ARDS (acute respiratory distress syndrome), and sepsis. TECO consistently showed better performance and was able to predict death risk earlier than other methods. Additionally, TECO identified key health indicators associated with ICU mortality, making its predictions more interpretable for clinicians. These findings suggest that TECO could become a valuable early warning tool, helping doctors monitor patients’ health and take timely action in a range of critical care situations.

## BACKGROUND AND SIGNIFICANCE

Modern medical and information technologies increasingly produce massive amounts of electronic health record (EHR) data. However, there still exists a gap between the rapid digitization of healthcare and the development of analytic tools capable of informing and guiding real-world clinical practices.[1, 2] Intensive care unit (ICU) is one area ripe for improved predictive analytics.[3] The COVID-19 pandemic posed unprecedented challenges to the provision of ICU care due to a lack of bed capacity, clinical care staff, and necessary analytics for resource allocation.[4] While the contemporary ICU medical devices and systems collect vast amounts of time-stamped data (e.g., vital signs, lab tests, and medication administrations), these rich continuous data streams are often not used to their full extent to develop analytics tools that assist clinicians to predict clinical outcomes.

One reason for this failure has been the analytics applied to such data. Conventional statistical models are limited in processing multivariate, time-dependent datasets and analyzing relations between different variables and different timestamps. Commercial analytics tools often lack technical transparency and interoperability across EHR platforms. For instance, the Epic Deterioration Index (EDI)[5, 6], a proprietary machine learning-based metric only available on Epic systems, was designed to quantify the level of deterioration patients experience at a point in time. The EDI utilizes data such as age, vital signs, laboratory tests, yet without considering patients’ overall comorbidities.[7, 8] The detailed design and parameters of EDI’s model have not been publicized, and EDI has not been validated in routine clinical practice.[8] Moreover, the threshold of EDI that calls for intervention has been differentially implemented among healthcare systems, rendering its actual usage dependent on local standards.[8]

As an alternative, machine learning models have been proposed to predict clinical outcomes or procedures in critically ill patients. For example, researchers employed a random forest (RF) classifier to predict COVID-19 disease severity at hospital admission,[9] applied an extreme gradient boosting (XGBoost) based algorithm to predict invasive mechanical ventilation,[10] and implemented PICTURE (Predicting Intensive Care Transfers and Other Unforeseen Events) to predict deterioration.[11] However, these algorithms face limitations when dealing with long series of time-dependent data with high dimensionality and irregular time intervals, as commonly encountered in inpatient monitoring data.

Recent advancements in deep learning have demonstrated preliminary success in managing multi-dimensional sequential EHR data, effectively capturing complex temporal patterns and interrelated features. Among these advancements, transformer models have shown promise by leveraging attention-based mechanisms to enhance both performance and efficiency in clinical settings.[12] Transformers employ multi-head attention and skip connections, removing the recurrent dependencies characteristic of earlier architectures like recurrent neural networks (RNNs) and long short-term memory networks (LSTMs),[13] while using positional embeddings to encode temporal information directly. Transformer-based architectures have since been adapted for various clinical applications. For example, Wu et al. developed a transformer model to forecast influenza prevalence from public health data, [14] while ClinicalBERT predicts 30-day hospital readmissions from clinical notes.[15] Models such as BEHRT [16] and Med-BERT [17], pre-trained on sequences of diagnosis codes, have been used to predict future diagnoses, with Antikainen et al. expanding this approach to incorporate different types of medical events for long-term mortality prediction in cardiovascular patients. [18]

In an ICU setting, MeTra integrates chest radiographs with clinical data for mortality prediction, though it restricts data input to the first 48 hours and may pose a high computational burden due to the Vision Transformer component.[19] Similarly, Cheng et al. used transformers for image data to predict COVID-19 mortality, but did not apply transformers to the clinical data component of their model.[20] Song et al. proposed the SAnD architecture for ICU tasks, including mortality prediction; however, they restricted data input to the final 24 hours of the ICU stay, requiring prior knowledge of the outcome time, which limits the model’s applicability to real-world scenarios.[21] Furthermore, none of these ICU mortality prediction models have been benchmarked against the widely-used commercial tool EDI, which limits the assessment of their clinical utility and relevance as a trans-platform tool.

## OBJECTIVE

In this study, we propose the Transformer-based Encounter-level Clinical Outcome (TECO) model, which fully utilizes continuous ICU monitoring data alongside patient-level baseline characteristics for mortality prediction after ICU admission, with the capability to update predictions on an hourly basis. We developed TECO using EHR data from a cohort of COVID-19 patients and validated the model on two external non-COVID-19 cohorts. We benchmarked TECO against the EDI, RF, and XGBoost to evaluate its performance in an ICU setting across different disease profiles.

## MATERIALS AND METHODS

### Study setting and design

In this study, the model development dataset contained EHR data from Texas Health Resources (THR), a large faith-based, nonprofit health system in North Texas, operating 20 acute care hospitals and serving 7 million residents in 16 counties. The THR cohort included 2579 adult patients with laboratory-confirmed COVID-19 (age ≥ 18), who were admitted into ICUs during their first COVID-19 hospitalization. Dates of patient hospitalization ranged from March 3, 2020 to August 13, 2021. Patients who had more than one ICU admission during the hospitalization were excluded.

The external validation dataset was extracted from the Medical Information Mart for Intensive Care (MIMIC-IV), a large, publicly available, de-identified clinical database for critically ill patients admitted to the emergency department of the Beth Israel Deaconess Medical Center in Boston, MA from 2008 to 2019.[22] We identified two patient cohorts admitted to ICU, one diagnosed with acute respiratory distress syndrome (ARDS) and the other with sepsis. If a patient had multiple ICU or hospital admissions, only the first ICU visit of the first hospital admission was included. ARDS was defined in accordance to the Berlin definition[23] with the MIMIC-specific positive identification method.[24] Sepsis was defined in accordance with the Third International Consensus Definitions for Sepsis and Septic Shock,[25] quantified by the Sequential Organ Failure Assessment (SOFA) score.[26, 27] Detailed definitions and query methods for identifying these two cohorts in the MIMIC-IV database are provided in the Supplementary Materials.

In this study, the models were designed to predict a binary outcome: death versus non-death in the ICU. In this context, the non-death outcome refers to conditions not at immediate risk of death and includes a range of states, from requiring continued ICU care to being ready for ICU discharge. We included two types of variables in the model: baseline variables and time-dependent variables. Baseline variables include age at hospital admission, sex, ethnicity, and race. Time-dependent variables include ICU monitoring measures, each recorded at a different and irregular pace: body temperature, respiration rate, pulse oximetry (SpO2), mSOFA (modified Sequential Organ Failure Assessment) overall score,[28] mSOFA respiratory sub-score, and SF ratio (SpO2/FiO2, where FiO2 is the fraction of inspired oxygen). Body mass index (BMI) was included as a time-dependent variable in the COVID-19 dataset. In the MIMIC-IV dataset, where time-dependent BMI was unavailable, the BMI recorded at hospital admission was used as a baseline variable.

The institutional review boards at THR and UT Southwestern Medical Center approved this study (Protocol #STU-2020-0786; activated on 8/24/2020). All patient identifiers were removed before EHR data extraction.

### Development of TECO Model

The TECO model employs a Transformer-encoder architecture. The overall algorithm design and data processing are illustrated in Figure 1. First, we aligned the time-dependent variables by taking the mean of each variable in every 15-minute interval. If a 15-minute mean value was missing, the value from the previous interval was carried forward. Then, we concatenated the aligned time-dependent variables with baseline variables, creating a feature set for each 15-minute interval, and embedded these features into a 512-dimension vector. The baseline variable input values remained static across the entire time range. The timestamps of these 15-minute intervals were encoded into another representative vector by positional encoding. We applied relative positional encoding to avoid imputing large amounts of default values for time points without measurement. The variable value vector and timestamp vector were fed into a feed-forward network with 6 Transformer-encoder layers. Each Transformer-encoder layer was equipped with 8 multi-head attention modules. The model outputs probability of death using a linear classification layer with the Softmax activation function.

**Figure 1.**
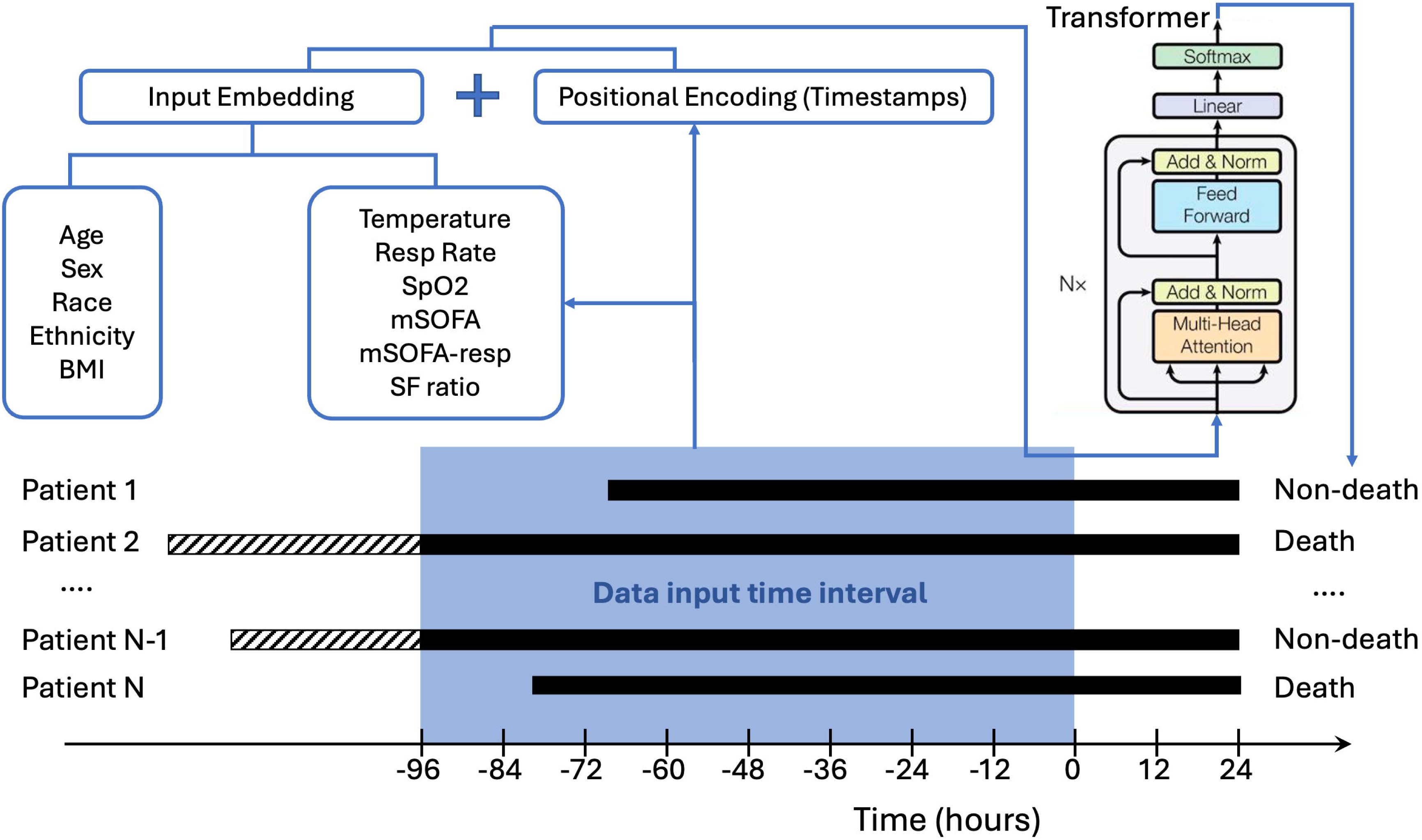
The TECO algorithm design. This figure demonstrates a 24-hour mortality prediction example, utilizing data from the preceding 96 hours to predict the binary outcome (death vs. non-death). Time-dependent, ICU monitoring variables were aligned and concatenated with baseline variables, then embedded into 512-dimensional feature vectors. These feature vectors were combined with positional timestamp vectors and fed into a multi-layer Transformer-encoder. (BMI: Body Mass Index. mSOFA: modified Sequential Organ Failure Assessment score. mSOFA-resp: mSOFA respiratory sub-score. Resp Rate: respiratory rate. SpO2: pulse oximetry, SF ratio: SpO2/FiO2 ratio. FiO2: fraction of inspired oxygen.)

We developed the TECO model using the THR COVID-19 dataset. We used the holdout method by creating 20 different data splits, where in each split the COVID-19 dataset was randomly split into a training set (80%) and a validation set (20%) on the patient level. During the training phase, we included only the data ranges that led to the eventual outcome at the ICU endpoint. This setup ensures a clear representation of both death and survival cases, while maintaining a balanced distribution between the two outcomes (36% death, Table 1). In this particular context, the non-death outcome corresponds to ICU discharge. In total, we trained nine TECO sub-models, each designed to predict the outcome at a specific future time point: 0, 12, 24, 36, 48, 60, 72, 84, and 96 hours. Each sub-model was provided with training data from a specific time interval (Supplementary Table 1). The algorithm does not require data to be fully available throughout the entire time interval. Records with incomplete time intervals were included in model training to ensure the model’s applicability in a clinical setting. To select hyperparameters, we performed a grid search on the hyperparameters on one training-validation split. The hyperparameter set yielding the highest area under the receiver operating characteristic (ROC) curve (AUC) from that split was selected, resulting in a model with 6 encoder layers, 8 attention heads, an embedding dimension of 512, and a total of 18,928,130 trainable parameters (Supplementary Table 2).

**Table 1.** Baseline characteristics in the COVID-19, ARDS, and sepsis cohorts.

|  | COVID-19 | MIMIC-ARDS | MIMIC-Sepsis |
| --- | --- | --- | --- |
| **Number of patients/encounters, n** | **2579** | **2799** | **6622** |
| **Age (years)** |  |  |  |
| Median (Q1, Q3) | 63.0 (51.0, 74.0) | 66.0 (54.0, 76.0) | 66.0 (54.0, 77.0) |
| **Sex, n (%)** |  |  |  |
| Male | 1499 (58.1) | 1595 (57.0) | 3782 (57.1) |
| *Female* | 1080 (41.9) | 1204 (43.0) | 2840 (42.9) |
| **Race, n (%)** |  |  |  |
| *White* | 1909 (74.0) | 1749 (62.5) | 4155 (62.7) |
| *Black* | 401 (15.5) | 217 (7.8) | 530 (8.0) |
| *Other* | 125 (4.8) | 168 (6.0) | 470 (7.1) |
| *Unknown* | 144 (5.6) | 665 (23.8) | 1467 (22.2) |
| **Ethnicity, n (%)** |  |  |  |
| *Hispanic* | 713 (27.6) | - | - |
| *Non-Hispanic* | 1723 (66.8) | - | - |
| *Unknown* | 143 (5.5) | - | - |
| **BMI, n (%)** |  |  |  |
| *Underweight* | 53 (2.1) | 53 (1.9) | 160 (2.4) |
| *Normal* | 379 (14.7) | 535 (19.1) | 1451 (21.9) |
| *Overweight* | 698 (27.1) | 652 (23.3) | 1673 (25.3) |
| *Obese* | 959 (37.2) | 792 (28.3) | 1540 (23.3) |
| *Unknown* | 490 (19.0) | 767 (27.4) | 1798 (27.2) |
| **Outcome, n (%)** |  |  |  |
| *Death* | 925 (35.9) | 471 (16.8) | 1031 (15.6) |
| *Discharge* | 1654 (64.1) | 2328 (83.2) | 5591 (84.4) |

(MIMIC: Medical Information Mart for Intensive Care. ARDS: acute respiratory distress syndrome.)

The Transformer model for developing TECO was implemented in PyTorch (Version 1.8.1)[29] and trained on an NVIDIA Tesla V100 Tensor Core GPU with 32 GB of memory. TECO was allowed to train for a maximum of 500 epochs with a batch size of 32. The SGD optimizer with momentum (0.9) was used to update model parameters.

The learning rate was set to 0.01 and reduced by a factor of 2 every 50 epochs. We set the dropout rate to 0 and use the Gaussian Error Linear Unit (GELU) as the activation function in the transformer layers. The training process would stop early if the validation loss did not change by more than 10^-4^ after 100 epochs. Gradient clipping was set to 1.0 to avoid gradient exploding (Supplementary Table 2).

### Development of Other Models for Comparison

To develop RF and XGBoost models using the same THR COVID-19 dataset, we followed the same data preparation procedure as described above for TECO. The time-dependent variables were aligned and averaged in 15-minute intervals, and concatenated with the baseline variables. The models were trained using the same 20 data splits as for TECO. Hyperparameters were selected through a grid search using the same one training-validation split as for TECO (Supplementary Table 2). RF and XGBoost were implemented in scikit-learn (Version 1.0.2).[30]

To use EDI for outcome prediction, we used the mean EDI value from the preceding 24 hours of each data input time interval. The EDI models predict the binary outcome solely based on a threshold on the continuous EDI values so no hyperparameter tuning was involved.

### Internal and External Validations

We evaluated the prediction performance of the TECO model based on AUC and compared it with the EDI, RF, and XGBoost models. Performance was evaluated separately for each of the nine TECO sub-models. For the internal validation based on the THR COVID-19 dataset, we reported the median AUCs on the validation sets across all 20 training-validation splits. To plot ROC for the EDI-based models, we used all possible thresholds within the range of the EDI data.

For external validation based on the MIMIC-IV dataset, we reported the AUCs and real-time probability of death on the ARDS and sepsis cohorts for each involved model, respectively. The EDI was evaluated only in the internal validation due to unavailability of EDI data in the non-Epic-based MIMIC-IV. To validate TECO externally in a manner more akin to a clinical setting, we positioned each model at varying time points after ICU admission and utilized a rolling window of the most recent available data to predict patient outcomes at future time points, as defined by each TECO sub-model’s task. For example, the 24-hour sub-model, which utilizes data from the preceding 96 hours (Figure 1), was employed to predict mortality at 120, 132, 144, and up to 240 hours after ICU admission (Figure 2).

**Figure 2.**
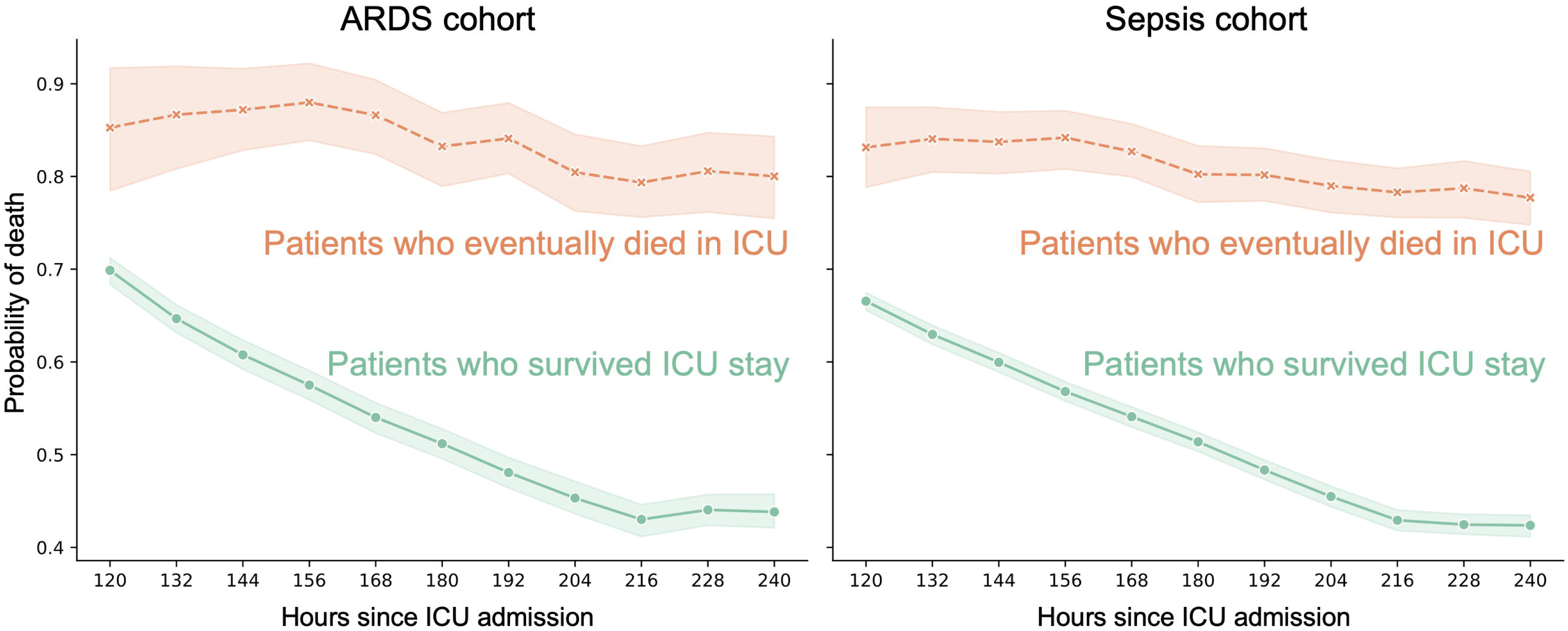
TECO-based monitoring of mortality probability in two external validation cohorts. The green line represents patients who were eventually discharged alive from ICU, while the orange line represents patients who died in ICU in the ARDS (left) cohort and sepsis (right) cohort, respectively. Probability of mortalities are aggregated over repeated hours since admission to show the mean and 95% confidence interval.

### Feature Importance and Ablation Study

To determine the feature importance of the ICU monitoring measures, we performed a feature elimination analysis on TECO with 20 random splits of training and validation. For each feature, the importance score was determined using the AUC information gained between the original model and the model without the feature. In addition, we calculated the impurity scores for RF and compared feature importance consistency between TECO and RF.

To assess the impact of baseline variables on model performance, we conducted an ablation study by comparing the AUCs of the full models to those of models using only time-dependent variables.

## RESULTS

A total of 2579 patients were included in the THR model development cohort. All enrolled patients had COVID-19, and of these, 925 (35.9%) expired in the ICU. The characteristics of the baseline variables are presented in Table 1. Among these patients, the median age was 63.0 years, and a majority were male (1499, 58.1%), white (1909, 74.0%), non-Hispanic (1723, 66.8%), and overweight to obese (BMI ≥ 25) (1657, 64.3%). The MIMIC ARDS validation cohort included 2799 patients, of whom 471 (16.8%) expired in the ICU. The MIMIC sepsis validation cohort included 6622 patients, of whom 1031 (15.6%) expired in the ICU. These two external validation cohorts presented similar trends in the distribution of baseline variables, with the majority being elderly, male, and white (Table 1).

### Internal Validation

In the COVID-19 model development cohort, AUCs on the validation sets across the 20 data splits are summarized in Supplementary Figure 1 and Supplementary Table 1. In general, all models’ performance improved as the targeted prediction time window was shortened, with the median AUCs ranging from 0.86 to 0.97. The median AUC of TECO model, ranging from 0.89 to 0.97, was higher than that of EDI (0.86–0.95), RF (0.87– 0.96), and XGBoost (0.88–0.96) at every prediction time window, demonstrating its overall advantages. On the other hand, the median AUC of EDI-based prediction was consistently lower than that from the other models at every prediction time window. It is noteworthy that the median AUC achieved by TECO when predicting 60-hour mortality (0.93) matched with that based on EDI when predicting 12-hour mortality (Supplementary Table 1), showcasing the advantage in early warning capability for TECO.

### External Validation

In the two external validation cohorts, similar trends of AUC were observed across different time intervals for each model (Table 2; Supplementary Figure 2). Using the sub-model for prediction of 24-hour mortality as a demonstration, for the ARDS cohort, AUCs for all three models improved from the earliest time point of prediction (120 hours since ICU admission), where the AUCs were 0.66 for TECO, 0.60 for RF, and 0.60 for XGBoost, to the latest time point (240 hours), where the AUCs were 0.76 (TECO), 0.73 (RF), and 0.72 (XGBoost), respectively. Similarly, for the sepsis cohort, AUCs of the same 24-hour sub-model improved from 0.65 (TECO), 0.57 (RF), and 0.57 (XGBoost) at the earliest prediction time point to 0.75 (TECO), 0.73 (RF), and 0.72 (XGBoost) at the latest time point (Table 2). Based on AUC, the TECO model consistently outperformed RF and XGBoost throughout a 5-day monitoring period in both cohorts. This advantage of TECO was particularly apparent at earlier lookout time points (e.g. 120 through 216 hours since ICU admission).

**Table 2.**
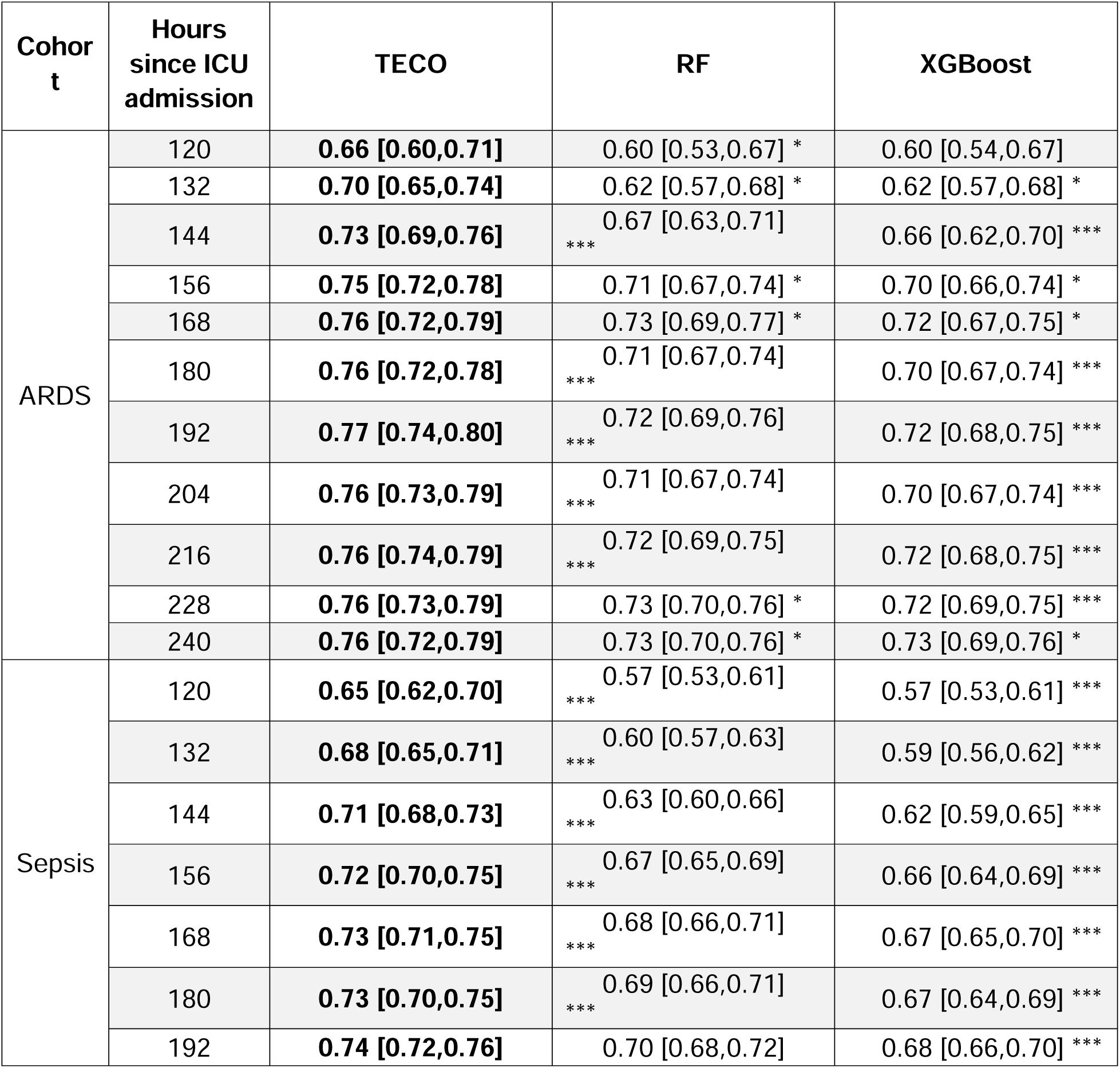
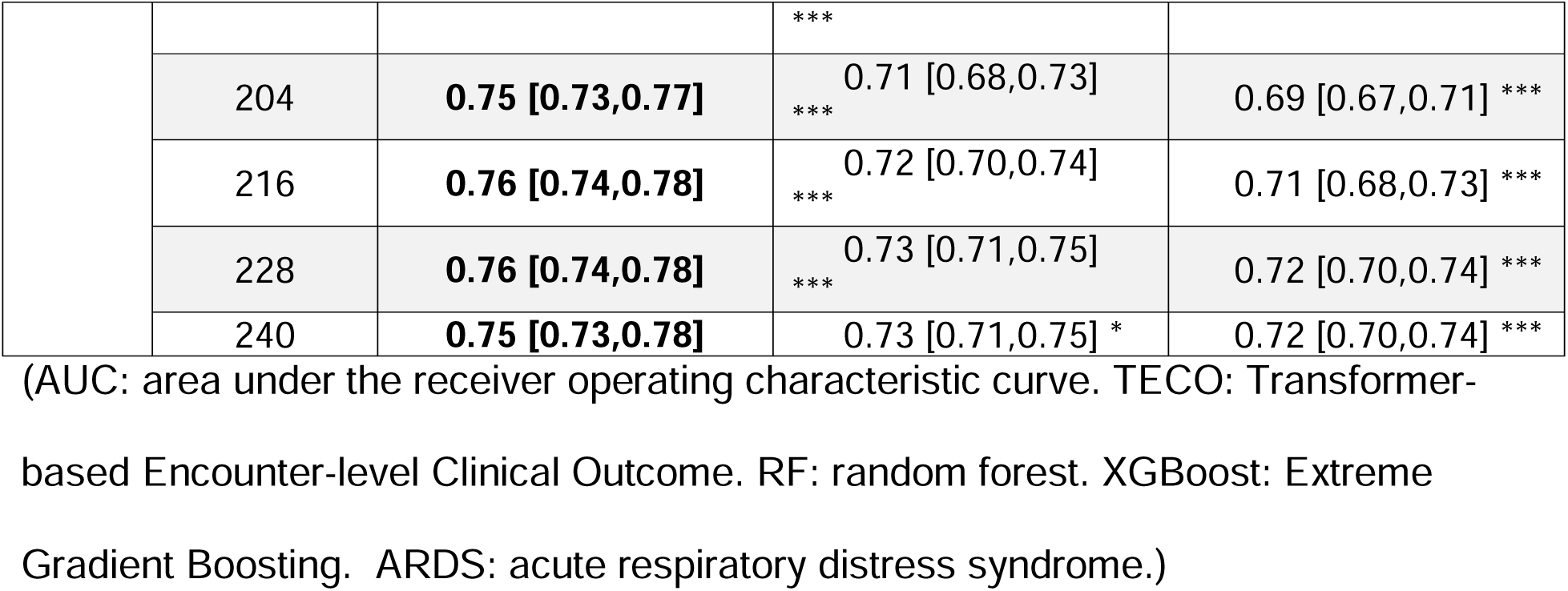
Model performance on the external validation cohorts to predict 24-hour mortality. At each time point after ICU admission, the models use the most recent 96 hours of data to predict mortality in the next 24 hours. For example, at 156 hours after ICU admission, the models use data from 60 to 156 hours to predict outcomes at 180 hours after admission. Model performances are presented as AUC [95% CI]. Confidence intervals (CIs) are estimated by bootstrapping with 500 iterations, sampling the whole dataset with replacement. Statistical significance was assessed using DeLong’s test to compare RF or XGBoost with TECO (* p < 0.05, *** p < 0.001).

As expected, the performance of all models in the external validation was generally lower than that in the internal validation. TECO’s performance in the ARDS cohort was slightly better than that in the sepsis cohort, especially at earlier lookout time points (Supplementary Figure 2).

### Monitoring of Patient Deterioration

TECO can be used to provide real-time estimation of mortality probability throughout the ICU stay. An illustration of this feature is shown in Figure 2, where results from external validations of the 24-hour sub-model are presented. TECO demonstrated that, at the cohort level, patients who ultimately survived their ICU stay exhibited a consistently lower probability of mortality throughout the 5-day monitoring period compared to those who eventually died in the ICU. Moreover, TECO displayed a decreasing trajectory in mortality probability for the surviving patients, a trend consistently observed across two external validation cohorts. Similar findings were observed with all nine TECO sub-models (Supplementary Figure 3). For an illustration of TECO-generated deterioration monitoring at the individual patient level, the mortality probability projection of eight representative patients from the ARDS and sepsis cohorts are shown in Supplementary Figure 4.

### Feature Importance and Ablation Study

In the feature importance analysis, SF ratio appeared to be the most important feature for TECO, especially at later time points closer to the outcome (Figure 3). The mSOFA demonstrated the second highest importance. Similarly, the mSOFA and SF ratio showed higher importance for RF, where the importance of SF ratio gradually increased towards the time of the outcome.

**Figure 3.**
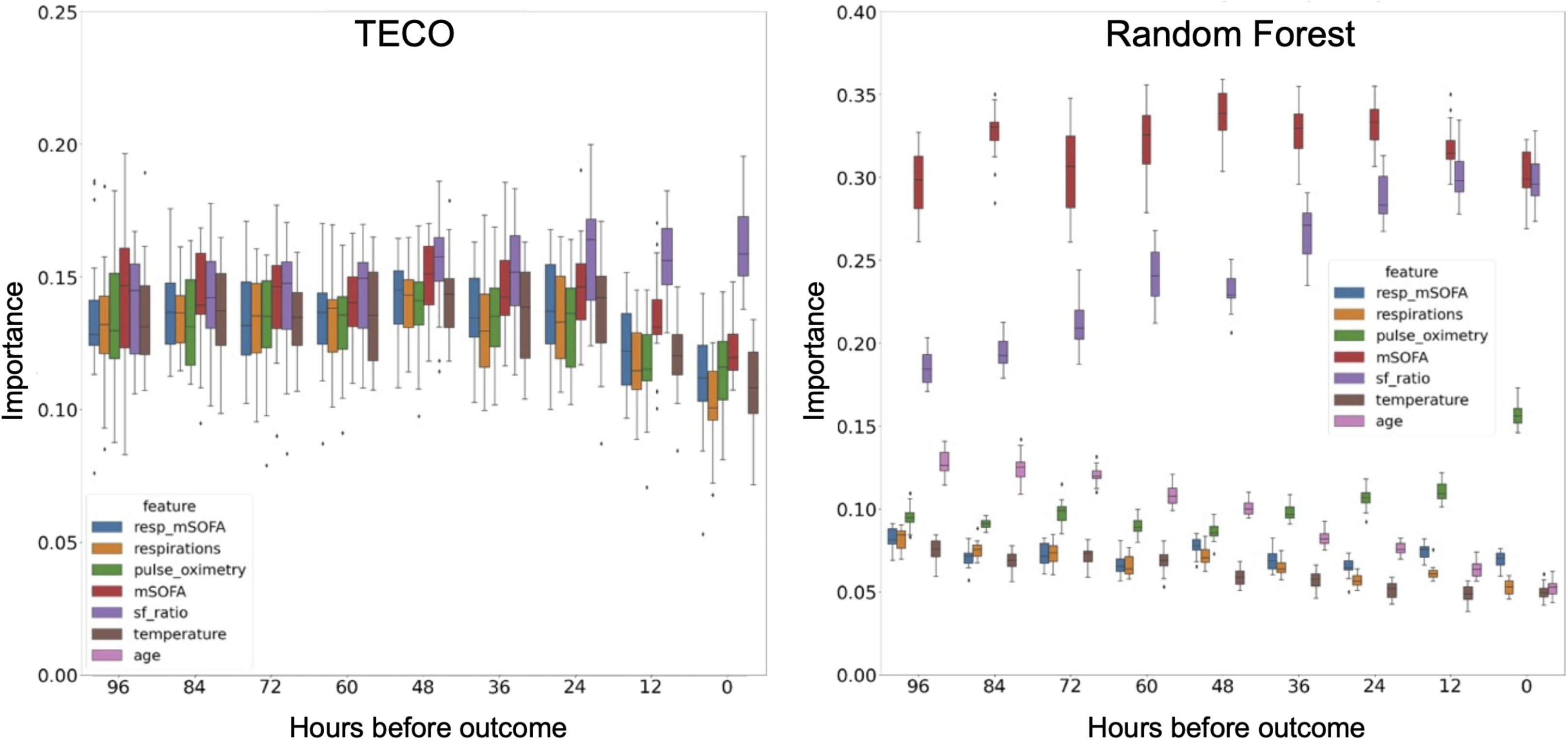
Feature importance analysis in the COVID-19 cohort. The left panel shows the information gain calculated through feature elimination for TECO. Each bar represents the AUC loss from 20 different validation splits. The right panel shows the random forest impurity importance for each feature with score > 0.05.

Removing the contribution of baseline variables led to a performance decrease across all models (TECO, RF, and XGBoost) in the two external validation cohorts (Supplementary Table 3). This performance drop was consistent across models and at different time points when the predictions were made. Notably, the contribution of these baseline variables appears to be independent of the timing of prediction. Importantly, even without baseline variables, TECO remained the top-performing model, particularly at earlier prediction time points, underscoring its intrinsic capability to effectively handle dynamic, time-dependent data.

## DISCUSSION

In this study, we developed and validated a novel deep learning algorithm, TECO, for mortality prediction in the ICU. Some existing methods for ICU mortality prediction also utilize transformer architecture and continuous monitoring data, particularly those available from the MIMIC-III and –IV databases.[19, 21] In contrast to these approaches, TECO is a lightweight transformer model specifically tailored to handle time-dependent, irregularly recorded features and time-independent baseline features in a joint manner. Unlike MeTra or Song et al. models, TECO does not presume a fixed time range of either the input data or the outcome. [19, 21] Instead, it can leverage the most recent ICU data to make predictions at future time points.

Our work benchmarks TECO against the commercially available EDI, a closed-source metric exclusive to Epic platforms. The robust performance of TECO, in comparison to EDI, highlights its potential as a better outcome prediction tool that is not confined to a single EHR system provider. The EDI was developed prior to the COVID-19 pandemic but was widely used for clinical decision support and ICU triage during the pandemic.[6, 8] In this study, we demonstrate that the EDI had relatively lower predictive performance beyond the 24-hour window prior to the outcome. In contrast, all three non-proprietary models—RF, XGBoost, and TECO—showed advantages especially at time points further from the ICU outcome. According to limited public information, the EDI model does not appear to use SF ratio in its development. [5, 6] Our feature importance analysis demonstrates that SF ratio could be of high importance, which may explain the limited performance of EDI. Besides SF ratio, mSOFA also had a high impact on the RF and TECO. This is consistent with the findings that SOFA is a reliable indicator for mortality among COVID-19 patients,[31–33] and that mSOFA has an equivalent performance in mortality prediction.[28]

Compared with RF and XGBoost in each external cohort, TECO outperformed the non-transformer models throughout a 5-day monitoring period, especially on the earlier days in the ICU. Calibration plots also reveal a stronger separation of outcomes based on TECO-estimated mortality probabilities (Supplementary Figure 5). TECO’s multi-head attention modules overcomes a bottleneck in traditional recurrent neural networks to learn long-range dependencies in sequences by linearly projecting the dimensions and queries of the input embedding.[12] The validation of TECO in these non-COVID-19 external cohorts underscores its generalizability in various severe diseases. It is also worth noting that TECO’s performance is comparable to several well-established ICU and in-hospital mortality prediction algorithms.[34–40] Some of these algorithms, while showing better predictive capabilities,[36, 37, 39], are limited by some essential factors such as a lack of external validation, a significantly smaller development sample size, challenges in real-time monitoring implementation, or a combination of these.

We demonstrate TECO’s practical utility in the ICU setting by highlighting its ability to monitor patient deterioration. The TECO-estimated mortality risk was consistently elevated among patients who eventually expired in the ICU, reflecting their heightened illness severity (Figure 2). To exclude the possibility that such trends were systemically introduced due to model artifacts, we examined and compared the mortality probability of individual patients (Supplementary Figure 4). Importantly, we observed different patterns among these patients, especially in the earlier days in ICU. Some of these patterns (e.g., Patients D, F, I) differ significantly from the aggregate trends observed at the group level (Figure 2), highlighting TECO’s ability to capture real-time, patient-specific details.

Developing an algorithm that can leverage the longitudinal time course of inpatient EHR data may improve prediction accuracy and enable earlier detection. In this study, the median AUC achieved by TECO using data up to 60 hours before the outcome (0.93) matched with that based on EDI at 12 hours before the outcome. With the successful validation of the external cohorts, this may suggest that TECO could signal a deterioration alert a full 48 hours before EDI. For ICUs with heavy workloads such as those observed during the COVID-19 pandemic, this improvement could substantially facilitate hospital resource planning, clinician communication with patient families, and play a vital role in future public health emergencies.

## LIMITATIONS AND FUTURE DIRECTIONS

This study has several limitations. Firstly, the usage of COVID-19 data to develop the TECO model was primarily motivated by the large sample size accumulated over the pandemic in our EHR systems. In addition, this Epic-based dataset included the proprietary EDI as a benchmark to evaluate our model—an option not available in other public ICU data sources. Ideally, external validation could have been conducted on a COVID-19 cohort from a different health system. Unfortunately, such data were not available at the time of this study. When validating TECO in the two external, non-COVID-19 cohorts, we found the model’s performance decreased compared with that in the COVID-19 cohort. While a performance drop from the training setting onto the independent testing setting is frequently observed [41, 42], it is important to note that the two external validation cohorts in this study represent two distinct diseases that differ from COVID-19 and the models were not trained on these diseases. Another potential contributor to the observed performance drop may stem from the fact that, when training TECO using the COVID-19 cohort, we only utilized patient outcomes at the ICU endpoint (i.e., death or discharge). This approach was intended to ensure representation of the two extreme scenarios across a broad spectrum of patients’ physiological conditions in the ICU. However, when evaluating TECO in the external cohorts over a moving time scale (120-240 hours since ICU admission), patients who are not at immediate risk of death may not necessarily present a health status that is ready for discharge. While TECO demonstrates proof of concept as a potential ICU monitoring tool, further validation across a broader range of disease states and ICU settings could significantly improve its performance and generalizability.

Nevertheless, our study demonstrates the feasibility of directly using structured EHR data, especially ICU monitoring data, to empower deep learning-based outcome prediction. The current version of TECO incorporates only 11 variables among which 6 are typically measured in the ICU in real time. Future work will need to consider whether more complex models using additional variables could influence the models’ performance and generalizability, especially given TECO’s intrinsic advantages in handling long-range, high-frequency data. However, operational costs associated with more complex and computationally intensive models must be thoroughly evaluated when considering these potential improvements. The TECO model in this study remains a lightweight transformer, which should not present significantly greater implementation challenges than the more conventional RF and XGBoost models. Moreover, with the advancement of large language models, clinical notes may also be used as additional features to provide valuable insights.[43–46]

Secondly, missing data and inconsistent data quality across different health systems or sites may significantly limit the applicability of a data-intensive model like TECO. For example, erroneous data due to instrument or human operations are frequently captured in the ICU data. More comprehensive data quality screening and control could benefit the implementation of TECO in real-world settings.

Lastly, it is worth noting that patients diagnosed with ARDS could potentially meet the sepsis criteria. Despite the absence of these cohorts in the training data for TECO, ensuring no information leakage, this scenario may still introduce a degree of bias and pose challenges to the implementation of TECO in the ICU and the interpretation of its outputs.

## CONCLUSION

We developed TECO, a transformer-based model, to analyze multi-dimensional, continuous monitoring data for ICU mortality prediction. In internal validation, TECO outperformed EDI-based prediction and other conventional machine learning methods. In two external validation cohorts (where EDI was not available), TECO outperformed other conventional machine learning methods. TECO may be further tailored as a disease-generic early warning tool in the ICU or inpatient settings.

## DATA AVAILABILITY STATEMENT

The COVID-19 dataset could not be shared publicly due to data and privacy protection policies at Texas Health Resources and UT Southwestern Medical Center. The MIMIC dataset is publicly available at https://physionet.org/content/mimiciv/2.2/.

## AUTHOR CONTRIBUTIONS

Conceptualization: RR, ZG, GX, DMY, YX Data curation and formal analysis: RR, ZG, HL, TN, TK, CW, CC, FV, GX, DMY, YX Methodology: RR, ZG, CC, AMN, EDP, GX, DMY, YX Writing: RR, ZG, KWJ, CC, AMN, FV, EDP, GX, DMY, YX

