## supplemental files for "A deep learning model for clinical outcome prediction using longitudinal inpatient electronic health records"

### SUPPLEMENTARY MATERIALS

**Formation Criteria of the External Cohorts**

The definition of the sepsis cohort was based on the MIMIC Code Repository (https://github.com/MIT-LCP/mimic-code/blob/main/mimic-iv/concepts/sepsis/sepsis3.sql). In the MIMIC-IV database, we identified the sepsis cohort by cross-referencing the Sequential Organ Failure Assessment (SOFA) and suspected infection tables to identify the earliest time when a patient's SOFA score was greater or equal to 2, coupled with evidence of a suspected infection

The definition of the ARDS cohort was based on Yang et al.'s procedure. [1]
In the MIMIC-IV database, we utilized the ICU stay, ventilation, and first-day blood gas tables to identify patients with an ICU stay exceeding two days and a PaO2/FiO2 ratio below 300.

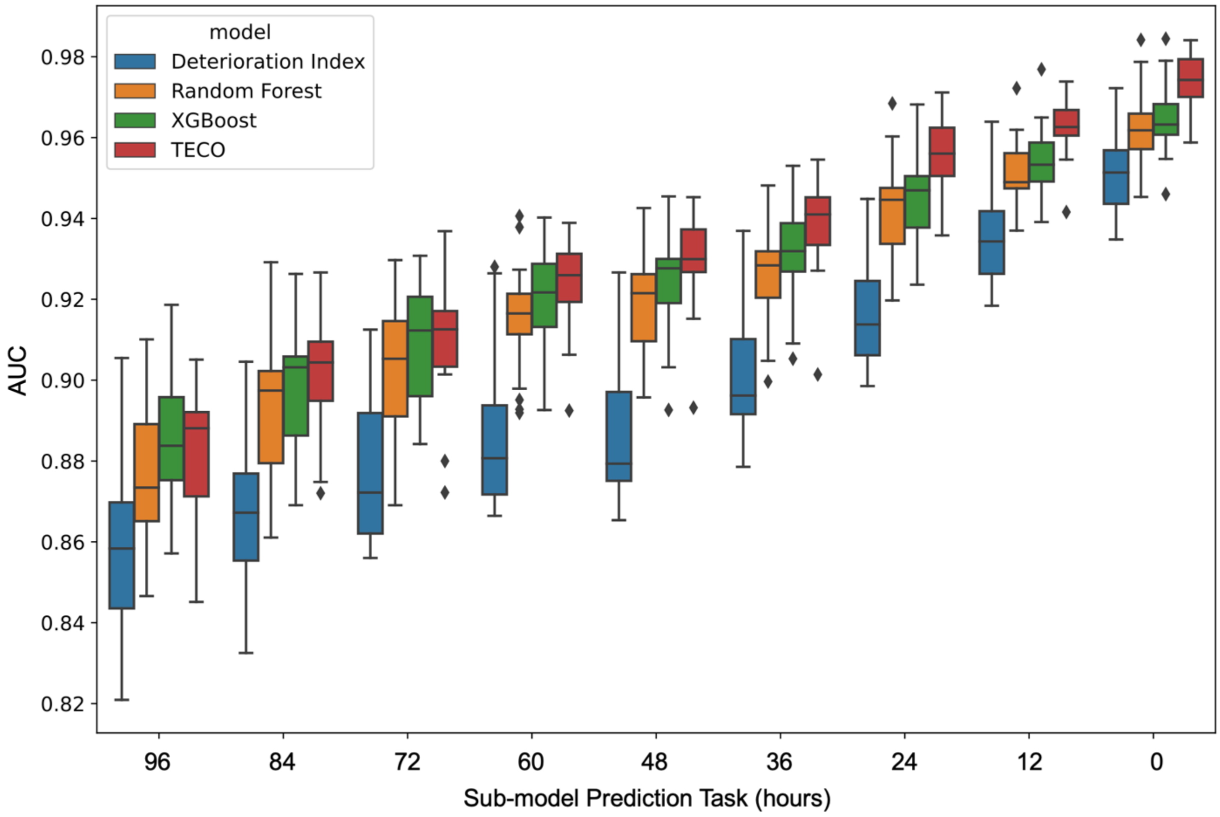


**Supplementary Figure 1. AUCs on the validation sets of the 20 data splits in the COVID-19 cohort.** AUC: area under the receiver operating characteristic curve. Deterioration Index: Epic Deterioration Index. XGBoost: eXtreme Gradient Boosting. TECO: Transformer-based Encounter-level Clinical Outcome.

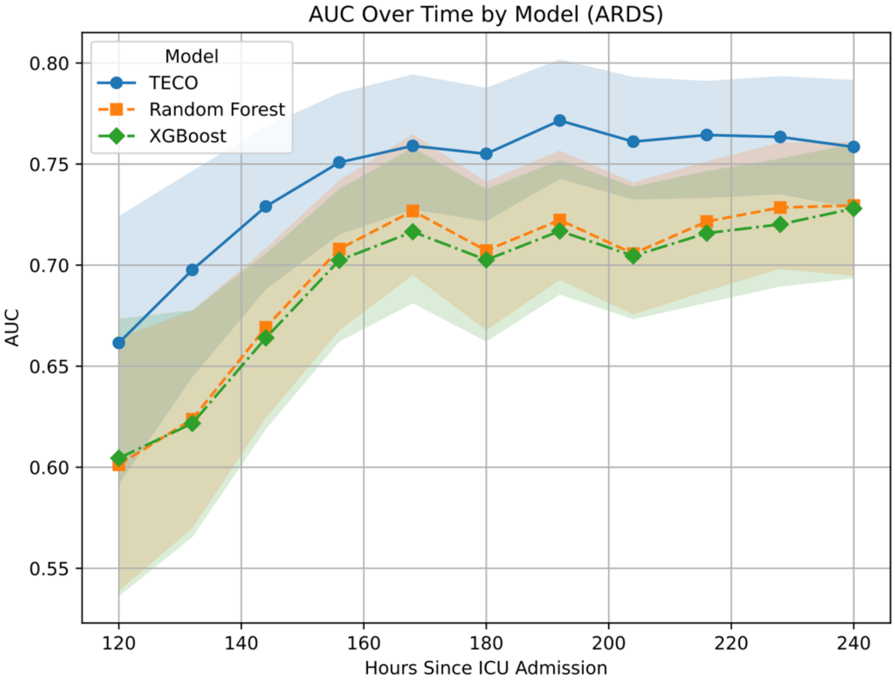


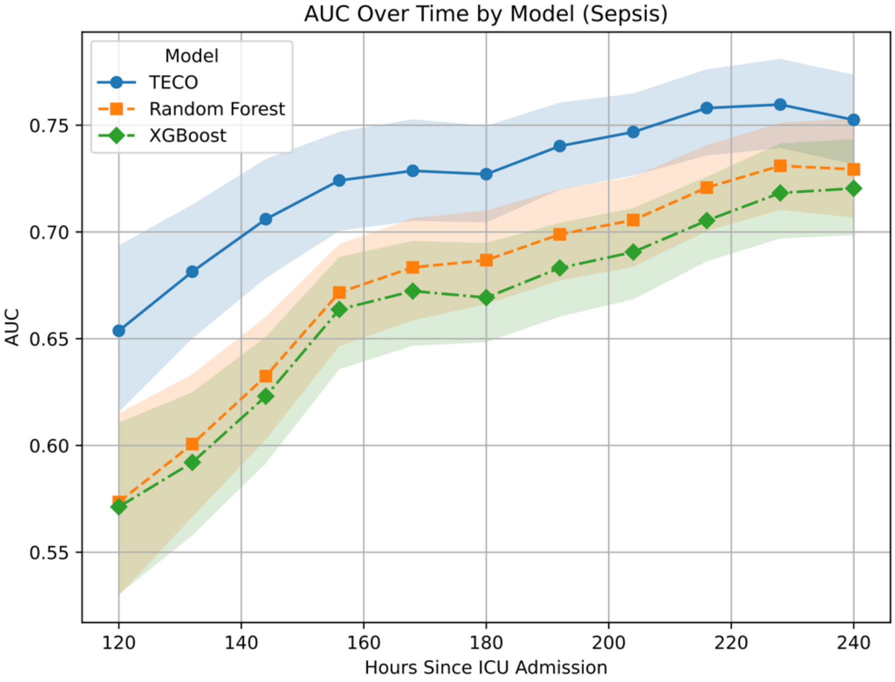


**Supplementary Figure 2. AUCs on the ARDS and sepsis cohorts.** AUC: area under the receiver operating characteristic curve. ARDS: acute respiratory distress syndrome. XGBoost: Extreme Gradient Boosting. TECO: Transformer-based Encounter-level Clinical Outcome.

a)

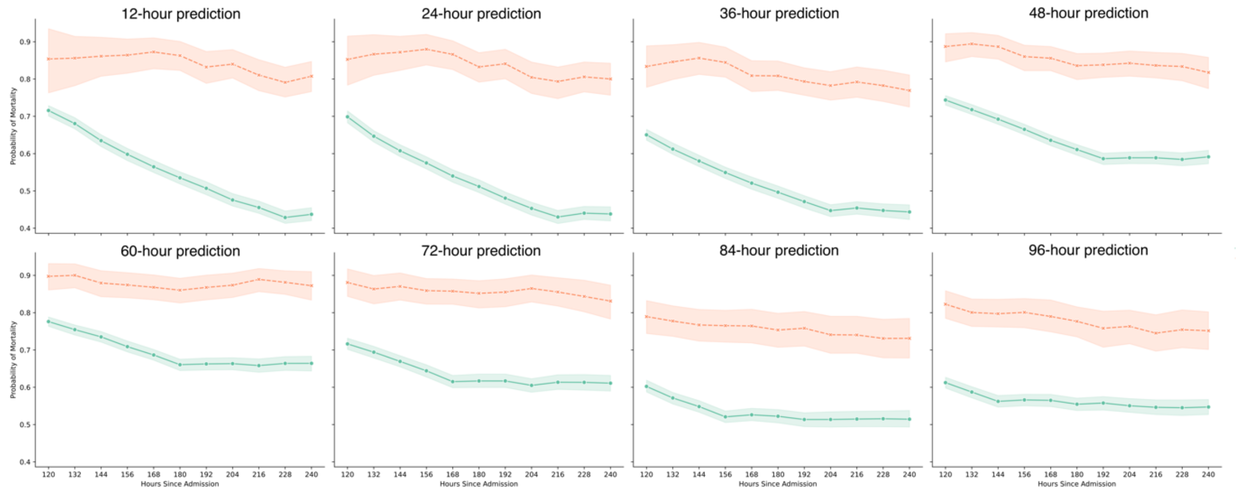


b)

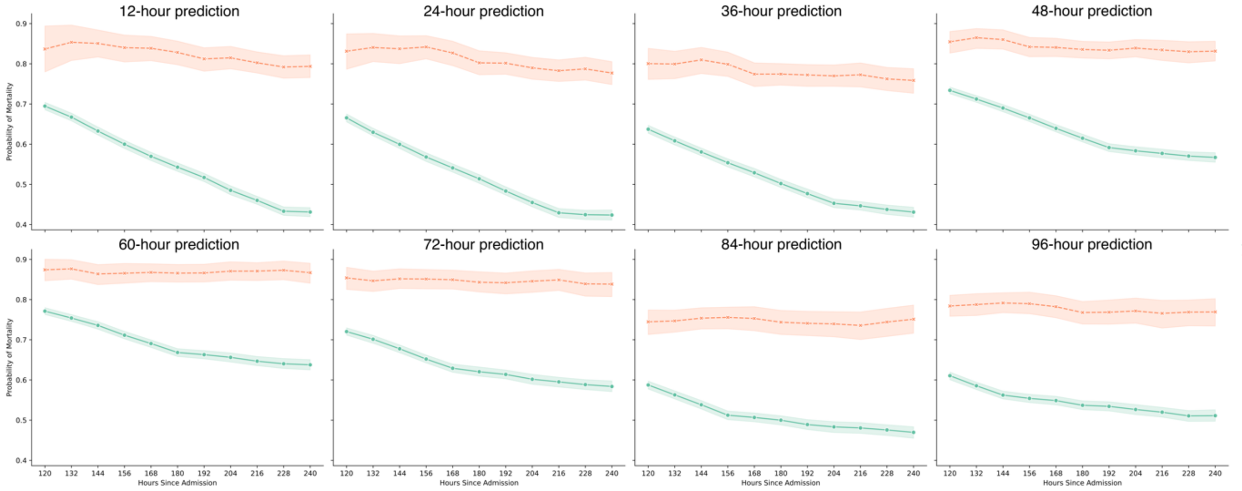


**Supplementary Figure 3. Prediction of mortality probability by nine TECO sub-models throughout a 5-day monitoring period in the a) ARDS cohort and b) sepsis cohort.** In x-axis, each time point indicates a lookout time of the model. The green lines represent patients who were eventually discharged alive from ICU, while the orange lines represent patients who died in ICU. Probability of mortalities are aggregated over repeated hours since admission to show the mean and 95% confidence interval.

**a)**

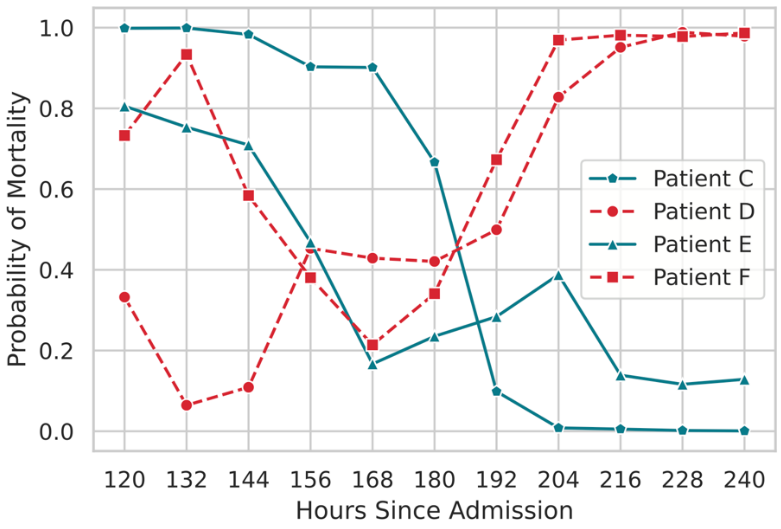


**b)**

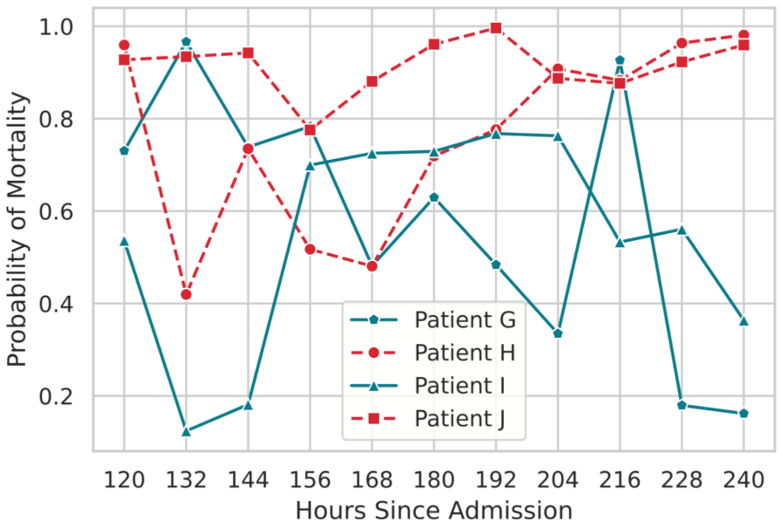


**Supplementary Figure 4. Comparison of TECO-based mortality probability over time between representative patients in a) ARDS cohort and b) sepsis cohort.** The green, solid line represents patients who survived in ICU, while the red, dashed line represents patients who died in the ICU, respectively.

**a) TECO**

**b) RF**

**c) XGBoost**

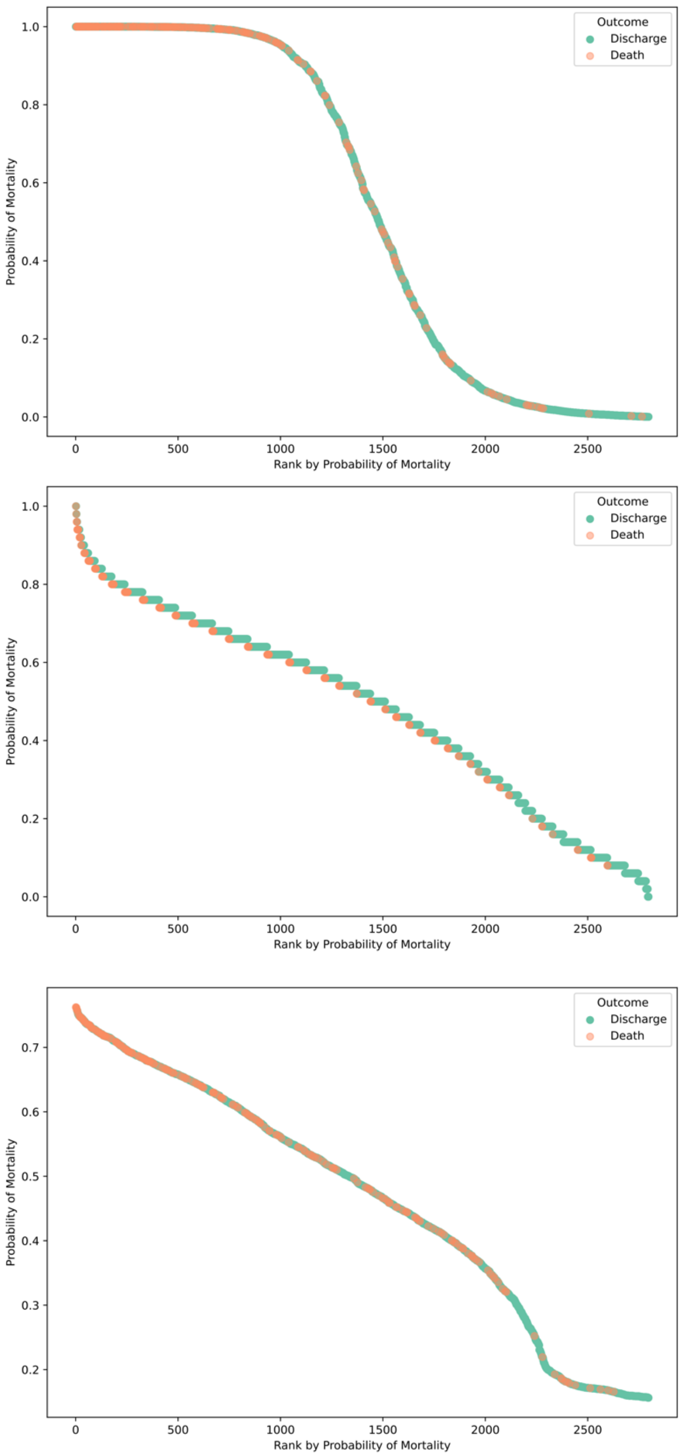


**Supplementary Figure 5. Calibration plots for models predicting 24-hour mortality.** These representative calibration plots were derived from the ARDS cohort, evaluated at 120 hours after ICU admission.

**Supplementary Table 1. Performance on the validation sets of the 20 data splits in the COVID-19 cohort.**

| Sub-model Prediction Task | Data Input Time Interval | Model | AUC | | |
|---|---|---|---|---|---|
| | | | Median | Q1 | Q3 |
| 96-hour mortality | Last 24 hours | TECO | 0.89 | 0.87 | 0.89 |
| | | EDI | 0.86 | 0.84 | 0.87 |
| | | RF | 0.87 | 0.87 | 0.89 |
| | | XGBoost | 0.88 | 0.88 | 0.90 |
| 84-hour mortality | Last 36 hours | TECO | 0.90 | 0.89 | 0.91 |
| | | EDI | 0.87 | 0.86 | 0.88 |
| | | RF | 0.90 | 0.88 | 0.90 |
| | | XGBoost | 0.90 | 0.89 | 0.91 |
| 72-hour mortality | Last 48 hours | TECO | 0.91 | 0.90 | 0.92 |
| | | EDI | 0.87 | 0.86 | 0.89 |
| | | RF | 0.91 | 0.89 | 0.91 |
| | | XGBoost | 0.91 | 0.90 | 0.92 |
| 60-hour mortality | Last 60 hours | TECO | 0.93 | 0.92 | 0.93 |
| | | EDI | 0.88 | 0.87 | 0.89 |
| | | RF | 0.92 | 0.91 | 0.92 |
| | | XGBoost | 0.92 | 0.91 | 0.93 |
| 48-hour mortality | Last 72 hours | TECO | 0.93 | 0.93 | 0.94 |
| | | EDI | 0.88 | 0.88 | 0.90 |
| | | RF | 0.92 | 0.91 | 0.93 |
| | | XGBoost | 0.93 | 0.92 | 0.93 |
| 36-hour mortality | Last 84 hours | TECO | 0.94 | 0.93 | 0.95 |
| | | EDI | 0.90 | 0.89 | 0.91 |
| | | RF | 0.93 | 0.92 | 0.93 |
| | | XGBoost | 0.93 | 0.93 | 0.94 |
| 24-hour mortality | Last 96 hours | TECO | 0.96 | 0.95 | 0.96 |
| | | EDI | 0.91 | 0.91 | 0.92 |
| | | RF | 0.94 | 0.93 | 0.95 |
| | | XGBoost | 0.95 | 0.94 | 0.95 |
| 12-hour mortality | Last 108 hours | TECO | 0.96 | 0.96 | 0.97 |
| | | EDI | 0.93 | 0.93 | 0.94 |
| | | RF | 0.95 | 0.95 | 0.96 |
| | | XGBoost | 0.95 | 0.95 | 0.96 |
| 0-hour mortality | Last 120 hours | TECO | 0.97 | 0.97 | 0.98 |
| | | EDI | 0.95 | 0.94 | 0.96 |
| | | RF | 0.96 | 0.96 | 0.97 |

| | | | | | |
|---|---|---|---|---|---|
| | | XGBoost | 0.96 | 0.96 | 0.97 |

AUC: area under the receiver operating characteristic curve. TECO: Transformer-based Encounter-level Clinical Outcome. EDI: Epic Deterioration Index. RF: Random Forest. XGBoost: eXtreme Gradient Boosting. Q1: first quartile. Q3: third quartile.

**Supplementary Table 2. Model hyperparameter grid-search and selection for TECO, random forests and XGBoost.**

| | Hyperparameter Options | Hyperparameter Selection | Description |
|---|---|---|---|
| **TECO** | d_model: [256, 512] | 512 | The embedding dimension |
| | nhead: [4, 8] | 8 | The number of heads in the multiheadattention models |
| | num_encoder_layers: [3, 6] | 6 | The number of layers in TransformerEncoder |
| | dim_feedforward: [1024, 2048] | 2048 | The dimension of the feedforward layer for classification |
| | dropout: [0, 0.1] | 0 | The dropout rate in the network |
| | activation: [‘gelu’, ‘relu’] | gelu | The activation function used in transformer layers |
| | optimizer: ['sgd', ('lr': 0.01, 'momentum': 0.9), ‘adamw’, ('lr': 1e-3, 'betas': (0.9, 0.999))] | 'sgd', ('lr': 0.01, 'momentum': 0.9) | The optimizer, learning rate and optimizer parameters. |
| | Gradient clipnorm: [0.5, 1.0, None] | 1.0 | Gradient norm scaling threshold. |
| **RF** | n_estimators: [50, 200, 400, 800] | 50 | The number of trees in the forest. |
| | max_depth: [20, 40, 80, None] | 20 | The maximum depth of the tree. |
| | max_features: [sqrt, auto] | auto | The number of features to consider when looking for the best split. |
| | min_samples_split: [2, 5, 10] | 2 | The minimum number of samples required to split an internal node. |
| | min_samples_leaf: [1, 2, 4] | 1 | The minimum number of samples required to be at a leaf node. |
| | bootstrap: [True, False] | True | bootstrap samples are used when building trees |
| **XGBoost** | loss: ['deviance'] | deviance | The loss function to optimize (logistic regression). |
| | n_estimators: [100, 200, 500] | 100 | The number of boosting stages to perform. |
| | learning_rate: [0.01, 0.1] | 0.01 | The learning rate shrinks the contribution of each tree. |
| | max_depth: [3, 5, 7, 10] | [3, 5, 7, 10] | The maximum depth of the individual regression estimators. |
| | subsample: [0.5, 0.7, 1.0] | [0.5, 0.7, 1.0] | The fraction of samples to be used for fitting the individual base learners. (<1.0 results in Stochastic Gradient Boosting.) |
| | max_features: ['auto'] | auto | The number of features to consider when looking for the best split. ‘auto’ uses ‘sqrt’ |

TECO: Transformer-based Encounter-level Clinical Outcome. RF: Random Forest. XGBoost: eXtreme Gradient Boosting

**Supplementary Table 3. Ablation study on the impact of baseline variables on model performance (24-hour mortality prediction).** At each time point after ICU admission, the models utilize the most recent 96 hours of data to predict outcomes in the next 24 hours. For example, at 156 hours after ICU admission, the models use data from 60 to 156 hours to predict outcomes at 180 hours after admission. Model performance is measured by the area under the receiver operating characteristic curve. The ablation study compares the performance of models without the contribution of baseline variables (i.e., using time-dependent variables only) against that of the full models (i.e., using both baseline and time-dependent variables). The contribution of baseline variables was removed by setting categorical variables (sex, race, and ethnicity) to "unknown" and setting continuous variables (age) to the cohort median.

| Cohort | Hours since ICU admission | TECO | | Random forest | | XGBoost | |
|---|---|---|---|---|---|---|---|
| | | Full model | Without baseline | Full model | Without baseline | Full model | Without baseline |
| ARDS | 120 | 0.66 | 0.63 | 0.61 | 0.60 | 0.61 | 0.60 |
| | 132 | 0.70 | 0.67 | 0.61 | 0.62 | 0.62 | 0.62 |
| | 144 | 0.73 | 0.70 | 0.67 | 0.67 | 0.67 | 0.66 |
| | 156 | 0.75 | 0.73 | 0.71 | 0.71 | 0.71 | 0.70 |
| | 168 | 0.76 | 0.74 | 0.73 | 0.73 | 0.73 | 0.72 |
| | 180 | 0.76 | 0.73 | 0.72 | 0.71 | 0.71 | 0.70 |
| | 192 | 0.77 | 0.75 | 0.73 | 0.72 | 0.73 | 0.72 |
| | 204 | 0.76 | 0.74 | 0.72 | 0.71 | 0.72 | 0.70 |
| | 216 | 0.76 | 0.74 | 0.73 | 0.72 | 0.73 | 0.72 |
| | 228 | 0.76 | 0.75 | 0.75 | 0.73 | 0.74 | 0.72 |
| | 240 | 0.76 | 0.74 | 0.74 | 0.73 | 0.74 | 0.73 |
| Sepsis | 120 | 0.65 | 0.63 | 0.59 | 0.57 | 0.59 | 0.57 |
| | 132 | 0.68 | 0.65 | 0.62 | 0.60 | 0.61 | 0.59 |
| | 144 | 0.71 | 0.68 | 0.65 | 0.63 | 0.64 | 0.62 |
| | 156 | 0.72 | 0.70 | 0.69 | 0.67 | 0.68 | 0.66 |
| | 168 | 0.73 | 0.71 | 0.70 | 0.68 | 0.69 | 0.67 |
| | 180 | 0.73 | 0.71 | 0.70 | 0.69 | 0.69 | 0.67 |
| | 192 | 0.74 | 0.72 | 0.71 | 0.70 | 0.70 | 0.68 |
| | 204 | 0.75 | 0.73 | 0.72 | 0.71 | 0.71 | 0.69 |
| | 216 | 0.76 | 0.74 | 0.74 | 0.72 | 0.72 | 0.71 |
| | 228 | 0.76 | 0.74 | 0.75 | 0.73 | 0.73 | 0.72 |
| | 240 | 0.75 | 0.73 | 0.75 | 0.73 | 0.74 | 0.72 |

(XGBoost: Extreme Gradient Boosting. TECO: Transformer-based Encounter-level Clinical Outcome. ARDS: acute respiratory distress syndrome.)
